# Long-lasting cigarette smoking alterations in immune function occur in cannabis smokers, possibly rendering them vulnerable to smoking-related tumors in later life

**DOI:** 10.1101/2024.04.01.24305156

**Authors:** Steven Lehrer, Peter H. Rheinstein

**Affiliations:** Department of Radiation Oncology, Icahn School of Medicine at Mount Sinai, New York; Severn Health Solutions, Severna Park, Maryland

**Keywords:** cigarettes, smoking, marijuana, cytokines, immune function

## Abstract

**Background:** Active cigarette smoking leads to increased CXCL5 production. CXCL5 mediates the immune response by attracting immune cells to areas of inflammation. Elevated CXCL5 levels are associated with various inflammatory diseases and tumorigenesis. In addition, smoking is linked to an increase in the level of the cytokine CEACAM6 in the bloodstream of smokers. CEACAM6 is increased in pancreatic adenocarcinoma, breast cancer, non⍰small cell lung cancer, gastric cancer, colon cancer and other cancers and promotes tumor progression, invasion, and metastasis. Although cytokine secretion in the innate immune response returns to nonsmoker levels after quitting smoking, the effects on the adaptive response appear to persist for years or decades due to epigenetic memory. As a result, epigenetic changes induced by smoking may contribute to long-lasting alterations in immune function, including elevated CXCL5 and CEACAM6. The effects of cannabis smoking might be similar.

**Methods:** In the current study we used UK Biobank (UKB) data to assess the relationship of CXCL5, CEACAM6, and pulmonary function to cigarette and cannabis smoking. Our UK Biobank application was approved as UKB project 57245 (S.L., P.H.R.). Our analysis included all subjects with smoking and/or marijuana use data in the UK Biobank database. Circulating levels of CXCL5 and CEACAM6 were from UKB Olink data. Individual CXCL5 and CEACAM6 levels are NPX, Normalized Protein expression, Olink arbitrary unit in Log2 scale (Olink Proteomics AB, Uppsala, Sweden; http://www.olink.com).

**Results:** Current smokers and past smokers had elevated circulating levels of CXCL5 and CECAM6. In multivariate analysis, current, past, or no smoking history was significantly related to CXCL5 level and CECAM6 levels, independent of the effects of age, sex. Frequency of cannabis use had a similar effect. In multivariate analysis, frequency of cannabis use was significantly related to CXCL5 level and CECAM6 levels, independent of the effects of age, sex, and years between last cannabis use and enrollment in study.

**Conclusion:** we can confirm a previous report of epigenetic changes induced by cigarette smoking that may contribute to long-lasting alterations in immune function related to CXCL5 and CEACAM6. In addition, we have found that these same long-lasting smoking alterations in immune function related to CXCL5 and CEACAM6 occur in cannabis smokers, possibly rendering them vulnerable to smoking-related tumors in later life.

---

Smoking has a lasting impact on the immune system, including alterations in adaptive immune response cytokine production. Specifically, active smoking is associated with an upregulation of the bacterially induced inflammatory cytokine C-X-C motif chemokine ligand 5 (CXCL5) [1, 2].

CXCL5 is a chemokine involved in inflammation and immune responses. It plays a role in recruiting and activating neutrophils to sites of infection or injury. Elevated CXCL5 levels are associated with various inflammatory diseases and tumorigenesis [3].

CXCL5 mediates the immune response by attracting immune cells to areas of inflammation. In addition, smoking is linked to an increase in the level of the protein CEA cell adhesion molecule 6 (CEACAM6) in the bloodstream of smokers. CEACAM6 is increased in pancreatic adenocarcinoma, breast cancer, non⍰small cell lung cancer, gastric cancer, colon cancer and other cancers and promotes tumor progression, invasion, and metastasis [4].

Although cytokine secretion in the innate immune response returns to nonsmoker levels after quitting smoking, the effects on the adaptive response appear to persist for years or decades due to epigenetic memory. As a result, epigenetic changes induced by smoking may contribute to long-lasting alterations in immune function [5].

Cannabis smoke may have similar deleterious effects on the airways and lungs as cigarette smoke. For example, human airway epithelial cells exposed to cannabis smoke elaborate CXCL5 [6].

In the current study we used UK Biobank data to assess the relationship of CXCL5, CEACAM6, and pulmonary function to cigarette and cannabis smoking.

## Methods

The UK Biobank is a large prospective observational study comprising approximately 500,000 men and women (N = 229,134 men, N = 273,402 women), more than 90% white, aged 40–69 years at enrollment. Participants were recruited from across 22 centers located throughout England, Wales, and Scotland between 2006 and 2010 and continue to be longitudinally followed for capture of subsequent health events [7]. This methodology is like that of the Framingham Heart Study [8], with the exception that the UKB program collects postmortem samples, which Framingham did not.

Our UK Biobank application was approved as UKB project 57245 (S.L., P.H.R.). Our analysis included all subjects with smoking and/or marijuana use data in the UK Biobank database.

The subject was asked, “Have you taken cannabis (marijuana, grass, hash, ganja, blow, draw, skunk, weed, spliff, dope), even if it was a long time ago?” If the answer was *yes*, cannabis use was recorded in UKB data field 20454, maximum frequency of taking cannabis, question asked: “Considering when you were taking cannabis most regularly, how often did you take it?” Answers were 1, Less than once a month; 2, Once a month or more, but not every week; 3, Once a week or more, but not every day; 4, Every day. Subject was then asked (UKB data field 20455), “About how old were you when you last had cannabis?”

CXCL5 and CEACAM6 were from UKB Olink data. Individual CXCL5 and CEACAM6 levels are NPX, Normalized Protein expression, Olink’s arbitrary unit in Log2 scale (Olink Proteomics AB, Uppsala, Sweden; http://www.olink.com) [9, 10]. Pulmonary function is expressed as Inverted GLI 2012 z-score for FEV1/FVC ratio. Methods for deriving this score can be found in Gupta et al 2017 [11].

UK Biobank: has approval from the Northwest Multi-center Research Ethics Committee (MREC) to obtain and disseminate data and samples from the participants, and these ethical regulations cover the work in this study. Written informed consent was obtained from all participants. Details can be found at www.ukbiobank.ac.uk/ethics.

Statistical analysis was done with SPSS v 26, (IBM, New York).

## Results

Mean age of subjects was 57 ± 8.1 (mean ± SD). 54% were women, 46% were men. 98% were white British.

Figure 1A illustrates CXCL5 normalized protein expression (mean + standard error, NPX) versus smoking status. Current smokers had the highest expression (p < 0.001, one way anova). Figure 1B illustrates CEACAM6 normalized protein expression (NPX) versus smoking status. Current smokers had the highest expression (p < 0.001). Figure 1C illustrates CXCL5 normalized protein expression versus frequency taking cannabis. Daily users had the highest expression (p < 0.013). Figure 1D illustrates CEACAM6 normalized protein expression (NPX) versus frequency taking cannabis. Daily users had the highest expression (p < 0.001). Figure 1E shows smoking status versus FEV1/FVC z-score. Never smokers had highest score (p < 0.001). Figure 1F shows frequency taking cannabis versus FEV1/FVC z-score. Daily users had highest score (p < 0.001).

**Figure 1A.**
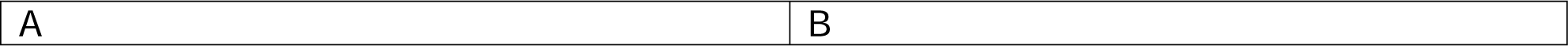
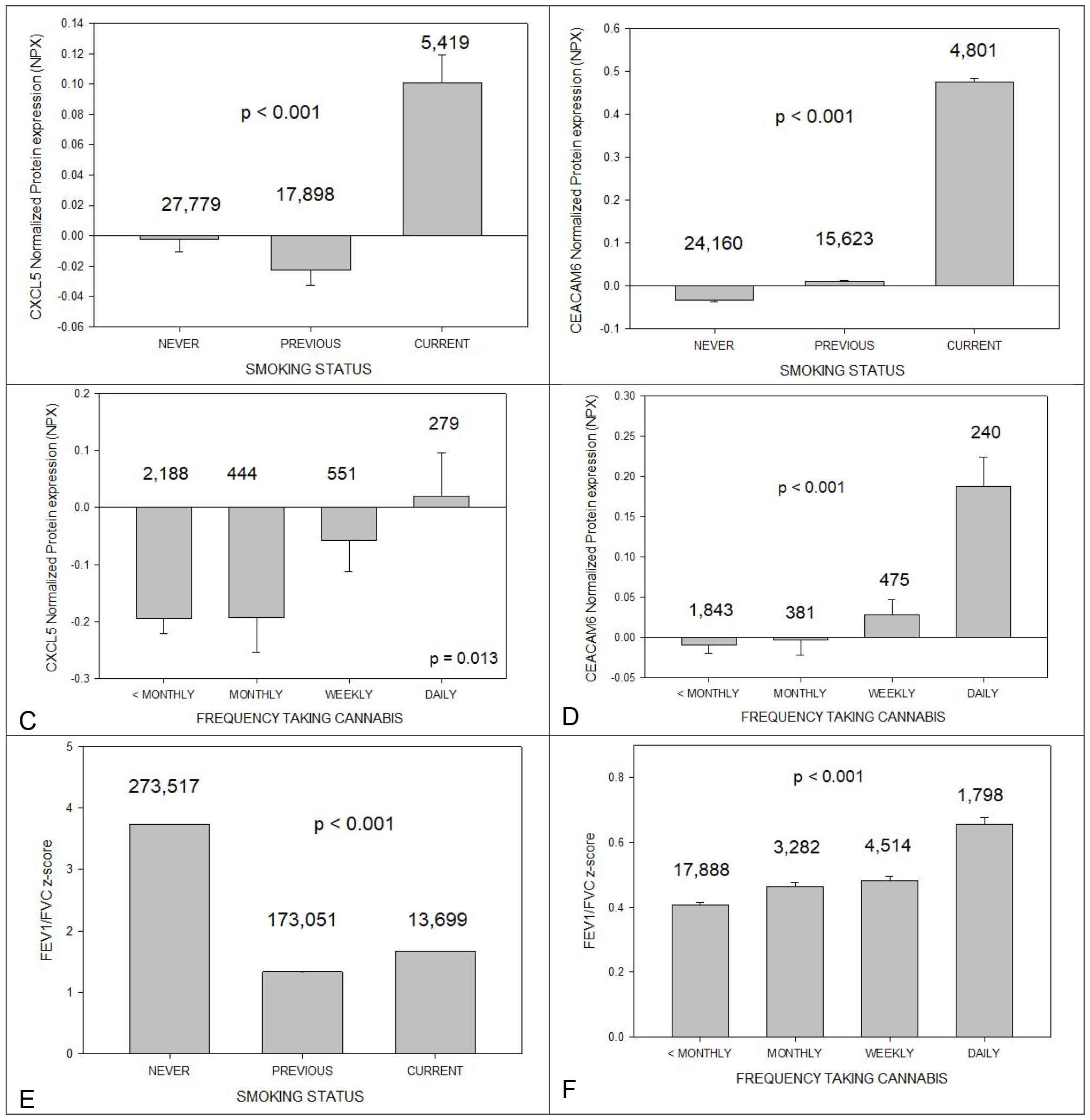
illustrates CXCL5 normalized protein expression (mean + standard error, NPX) versus smoking status. Current smokers had the highest expression (p < 0.001, one way anova). Number of cases in each group above corresponding bar. Figure 1B illustrates CEACAM6 normalized protein expression (NPX) versus smoking status. Current smokers had the highest expression (p < 0.001). Figure 1C illustrates CXCL5 normalized protein expression versus frequency taking cannabis. Daily users had the highest expression (p < 0.013). Figure 1D illustrates CEACAM6 normalized protein expression (NPX) versus frequency taking cannabis. Daily users had the highest expression (p < 0.001). Number of cases in each group above bar. Figure 1E shows smoking status versus FEV1/FVC z-score. Never smokers had highest score (p < 0.001). Figure 1F shows frequency taking cannabis versus FEV1/FVC z-score. Daily users had highest score (p < 0.001).

Figure 2 illustrates years between last cannabis use and enrollment in study (age gap) in 20996 subjects. On average, subjects had not smoked cannabis for 24 ± 11 years (mean ± S.D).

**Figure 2.**
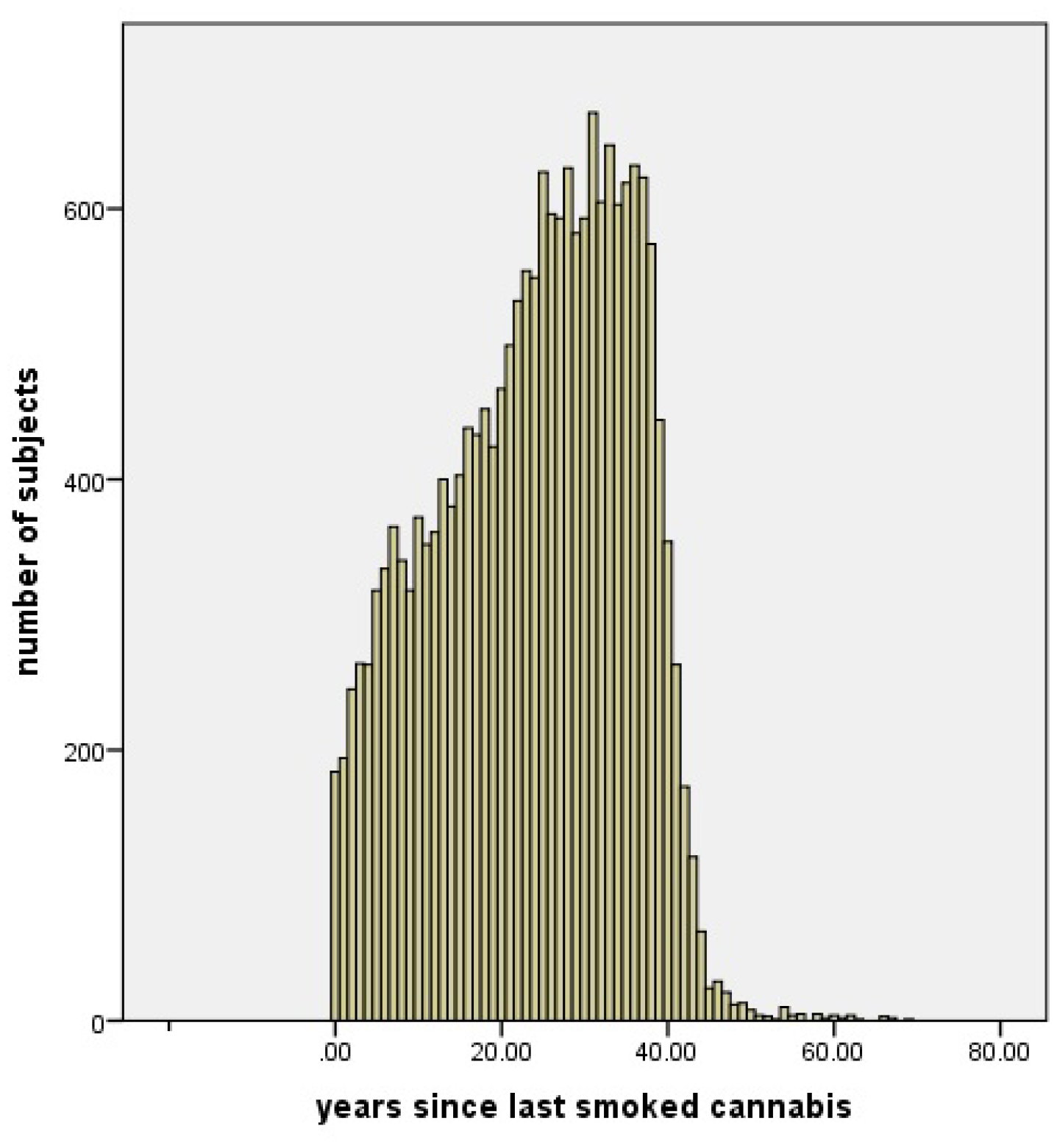
Years between last cannabis use and enrollment in study (age gap) in 20996 subjects. On average, subjects had not smoked cannabis for 24 ± 11 years (mean ± S.D).

To exclude the effects of age, sex, and smoking status (none, past smoker, current smoker), CXCL5 and CEACAM6 levels, multivariate linear regression was performed.

Table 1A shows multivariate linear regression, CXCL5 dependent variable, sex, age, cigarette smoking status (none, past, current). Women had significantly higher CXCL5 levels than men (p< 0.001), older subjects had significantly higher CXCL5 levels than younger subjects (p < 0.001), current and past smokers had higher CXCL5 levels than never smokers (p < 0.001).

**Table 1A.**
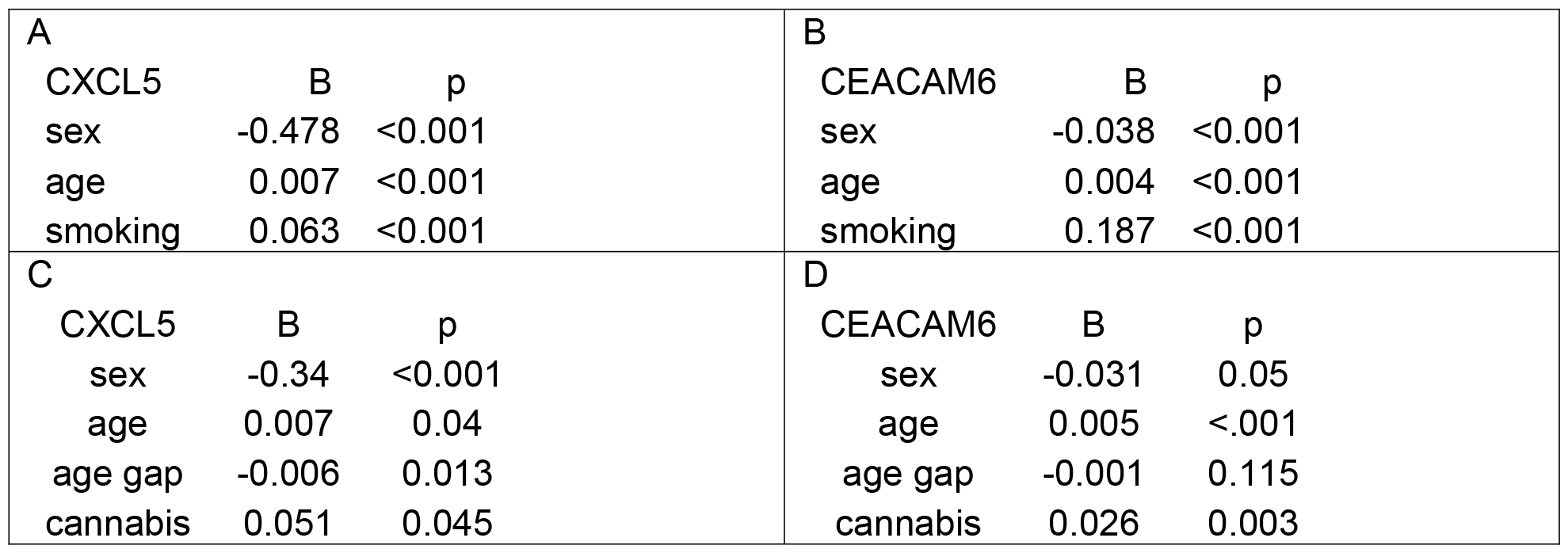
Multivariate linear regression, CXCL5 dependent variable, sex, age, smoking status (none, past, current). Women had significantly higher CXCL5 levels than men (p < 0.001), older subjects had significantly higher CXCL5 levels than younger subjects (p < 0.001), current and past smokers had higher CXCL5 levels than never smokers (p < 0.001). Table 1B. Multivariate linear regression, CEACAM6 dependent variable, sex, age, smoking status (none, past, current). Women had significantly higher CEACAM6 levels than men (p < 0.001), older subjects had significantly higher CEACAM6 levels than younger subjects (p < 0.001), current and past smokers had higher CEACAM6 levels than never smokers (p < 0.001). Table 1C. Multivariate linear regression, CXCL5 dependent variable, sex, age, age gap (Years between last cannabis use and enrollment in study), and frequency of cannabis use. Women had significantly higher CXCL5 levels than men (p < 0.001), older subjects had significantly higher CXCL5 levels than younger subjects (p = 0.04), lower age gap was significantly inversely related to higher CXCL5 (p = 0.013), and frequency of cannabis use was significantly related to CXCL5 level (p = 0.045). Table 1D. Multivariate linear regression, CEACAM6 dependent variable, sex, age, age gap (Years between last cannabis use and enrollment in study), and frequency of cannabis use. Women had significantly higher CEACAM6 levels than men (p = 0.05), older subjects had significantly higher CEACAM6 levels than younger subjects (p < 0.001), lower age gap was insignificantly inversely related to higher CEACAM6 (p = 0.115), and frequency of cannabis use was significantly related to CEACAM6 level (p = 0.003).

Table 1B shows multivariate linear regression, CEACAM6 dependent variable, sex, age, smoking status (none, past, current). Women had significantly higher CEACAM6 levels than men (p < 0.001), older subjects had significantly higher CEACAM6 levels than younger subjects (p < 0.001), current and past smokers had higher CEACAM6 levels than never smokers (p < 0.001).

To exclude the effects of age, sex, age gap (Years between last cannabis use and enrollment in study) on frequency of cannabis use versus CXCL5 and CEACAM6 levels, multivariate linear regression was performed.

Table 1C shows multivariate linear regression, CXCL5 dependent variable, sex, age, age gap (Years between last cannabis use and enrollment in study), and frequency of cannabis use. Women had significantly higher CXCL5 levels than men (p < 0.001), older subjects had significantly higher CXCL5 levels than younger subjects (p = 0.04), lower age gap was significantly inversely related to higher CXCL5 (p = 0.013), and frequency of cannabis use was significantly related to CXCL5 level (p = 0.045).

Table 1D shows Multivariate linear regression, CEACAM6 dependent variable, sex, age, age gap (Years between last cannabis use and enrollment in study), and frequency of cannabis use. Women had significantly higher CEACAM6 levels than men (p = 0.05), older subjects had significantly higher CEACAM6 levels than younger subjects (p < 0.001), lower age gap was insignificantly inversely related to higher CEACAM6 (p = 0.115), and frequency of cannabis use was significantly related to CEACAM6 level (p = 0.003).

The multivariate general linear model was calculated, CXCL5 and CEACAM6 dependent variables, smoking status and frequency of cannabis use, independent variables. Smoking status was independently related to CEACAM6 (p < 0.001) but not to CXCL5 (p = 0.462).

Frequency of cannabis use was independently related to CXCL5 (p = 0.031) but not to CEACAM6 (p = 0.190). These statistics suggest that in smokers who used cannabis, the effect of smoking was more important for CEACAM6 levels, while frequency of cannabis use was more important for CXCL5 levels.

## Discussion

FEV1/FVC decline in smokers and former smokers is well documented [12]. We found that decline is increased in former smokers compared to current smokers. We surmise that the reason is because former smokers had noted increased breathing difficulty and quit smoking. Trouble breathing had not yet bothered current smokers and so they were unmotivated to quit.

Our finding of increased FEV1/FVC with increased marijuana smoking is consistent with large studies which have shown that, instead of reducing forced expiratory volume in 1□s and forced vital capacity (FVC), marijuana smoking is associated with increased FVC. The cause of this is unclear; acute bronchodilator and anti-inflammatory effects of cannabis may be partly responsible [13]. But increased CXCL5 and CEACAM6 suggest that, as in cigarette smokers, immunologic epigenetic memory is retained, and cannabis smokers will be vulnerable to smoking related malignancies in later life. Multivariate analysis (Table 1) confirmed these findings.

In the case of lung cancer, for example, the latency period between the onset of cigarette smoking and the diagnosis of lung cancer can be quite variable. Some studies suggest that it may take several decades for lung cancer to develop after the initiation of smoking. Genomic changes in non-small cell lung cancer have indicated a long period of latency that precedes clinical detection [14]. Cannabis smoking may be similar.

Cannabis use has benefits and harms. Among the benefits, cannabis can suppress the immune system, which might be beneficial for certain infections and autoimmune diseases. For instance, cannabis could reduce the systemic inflammatory response associated with sepsis. Animal studies suggest that stimulating endocannabinoid receptors with cannabinoids can reduce infection-related inflammation and improve recovery in some cases [15]. Autoimmune diseases might also be treated with cannabis [16]. Although medical cannabis is not an FDA-approved treatment for cancer-related pain, many oncologists discuss and recommend it to their patients [17].

Among the harms of cannabis, US 12th-grade students’ self-reported prevalence of marijuana use over the past 12 months was 30.4% [18]; these young people may have increased vulnerability to smoking-related tumors in later life. Moreover, increasing cannabis use is associated with increased chronic obstructive pulmonary disease (COPD) prevalence and increased COVID-19 test positivity [19].

In conclusion, we can confirm a previous report of epigenetic changes induced by cigarette smoking that may contribute to long-lasting alterations in immune function related to CXCL5 and CEACAM6 [5]. In addition, we have found that these same long-lasting smoking alterations in immune function related to CXCL5 and CEACAM6 occur in cannabis smokers, possibly rendering them vulnerable to smoking-related tumors in later life.

## Data Availability

Data available after approved application to UK Biobank

https://www.ukbiobank.ac.uk/

## Notes

Conflicts of interest: The authors declare that they have no competing interests.

This work was supported in part through the computational and data resources and staff expertise provided by Scientific Computing and Data at the Icahn School of Medicine at Mount Sinai and supported by the Clinical and Translational Science Awards (CTSA) grant UL1TR004419 from the National Center for Advancing Translational Sciences.

### Competing Interest Statement

The authors have declared no competing interest.

### Funding Statement

This study did not receive any funding

### Author Declarations

Data from UK Biobank https://www.ukbiobank.ac.uk/

